# Assessing mpox knowledge and sexual behaviours within high-risk populations in the Democratic Republic of the Congo

**DOI:** 10.1101/2025.04.20.25326123

**Authors:** Candice Lemaille, Megan Halbrook, Sydney Merritt, Yvon Anta, Lygie Lunyanga, Patrick K. Mukadi, Emmanuel Hasivirwe Vakaniaki, Thierry Kalonji, Michel Kenye, Cris Kacita, Sylvie Linsuke, Isaac I. Bogoch, Muge Cevik, Gregg S. Gonsalves, Mikayla Hunter, Laurens Liesenborghs, Jean-Claude Makangara-Cigolo, Souradet Y. Shaw, Robert L. Shongo, Lisa E. Hensley, Nicole A. Hoff, Anne W. Rimoin, Placide Mbala-Kingebeni, Jason Kindrachuk

**Affiliations:** Department of Medical Microbiology and Infectious Diseases, University of Manitoba, Winnipeg, MB, Canada; Jonathan and Karin Fielding School of Public Health, Department of Epidemiology, University of California, Los Angeles, CA, USA; Institut National de Recherche Biomédicale (INRB), Kinshasa, Democratic Republic of the Congo; National Program for the Control of Mpox and Viral Hemorrhagic Fevers, Ministry of Health, Kinshasa, Democratic Republic of the Congo; Department of Medicine, University of Toronto, Toronto, ON, Canada; Division of Infection and Global Health, University of St Andrews, St Andrews, Scotland; Department of Epidemiology of Microbial Diseases, Yale School of Public Health, New Haven, CT, USA; Public Health Modeling Unit, Yale School of Public Health, New Haven, CT, USA; Department of Clinical Sciences, Institute of Tropical Medicine Antwerp, Antwerp, Belgium; Graduate School of Cellular and Biomedical Sciences, University of Bern, Bern, Switzerland; Department of Community Health Sciences, Max Rady College of Medicine, University of Manitoba, Winnipeg, MB, Canada; Agricultural Research Service, National Bio-/Agro-Facility, Zoonotic and Emerging Diseases Research Unit, United States Department of Agriculture (USDA), Manhattan, KS, USA; Department of Internal Medicine, University of Manitoba, Winnipeg, MB R3E 0W2, Canada; Manitoba Centre for Proteomics and Systems Biology, Health Science Centre, Winnipeg, Manitoba

## Abstract

**Background:** Historically, the Democratic Republic of the Congo (DRC) has faced the greatest public health burden from mpox, including more than 70,000 probable cases from 01 January 2024 to 02 February 2025. However, there has been a relative paucity of investigation focused on mpox community engagement in DRC, including assessments of disease knowledge and risk perception.

**Methods:** Given the ongoing Clade I mpox public health emergency of international concern, and the linkage between sustained human-to-human transmission and dense sexual networks, we sought to investigate mpox knowledge and sexual behaviours among key populations. Between March 20, 2024, and August 25, 2024, we recruited 2794 participants distributed across Kinshasa, Kwango and North Kivu provinces, with a focus in urban centers where mpox risk was considered high.

**Results:** Most participants were considered other at-risk populations (1035; 37.0%), followed by men who have sex with men (MSM, 831; 29.7%) and sex workers (810; 29.0%). Mpox knowledge, including transmission routes, as well as sexual and health-seeking behaviours were evaluated through questionnaires led by peer educators. Overall, only 6.1% of all participants reported prior mpox knowledge. Among this participant subset, zoonosis (“direct contact with infected animals”) and “people living in high-risk areas” were the most frequently selected options in regard to mpox transmission and populations at risk, respectively. When considering at-risk behaviors for mpox, those that identified as sex workers reported significantly higher risk sexual activities including multiple sexual partners (80.3% of sex work participants), engaging in transactional sex (84.7.0%), and anonymous sex (80.8%) compared to MSM. However, both sex workers (44.8%) and MSM (56.7%) reported the highest health seeking behaviors for a suspected sexually transmitted infection.

**Conclusion:** Our results highlight that community engagement which incorporates both mpox knowledge and risk perception activities and is inclusive of at-risk populations are needed for ongoing mpox containment and mitigation efforts.

## INTRODUCTION

In 2024, the World Health Organization (WHO) announced the second mpox public health emergency of international concern (PHEIC), two years following the first mpox PHEIC resulting from a global mpox outbreak that impacted over one hundred non-endemic countries [1, 2]. The Democratic Republic of the Congo (DRC), where Clade I mpox virus (previously known as monkeypox virus; MPXV) is endemic, has faced the largest public health burden from mpox globally. This includes more than 70,000 probable cases from 01 January 2024 to 02 February 2025 [3]. Historically, Clade I MPXV infections have been primarily driven through zoonotic contact with limited recorded secondary transmission events [4]. However, the 2023 emergence of Clade Ib MPXV in South Kivu, DRC, has resulted in shifting clinical and epidemiological characteristics. This has included sustained patterns of human-to-human transmission associated with sexual or intimate contacts and concentration among sexual networks [5, 6]. Additionally, extensive APOBEC3 mutations have also been identified among MPXV genomes associated with sustained human-to-human transmissions, similar to that seen among Clade IIb MPXV [7–10]. Sustained Clade Ia mpox transmission and APOBEC3 mutations have also been identified in Kinshasa. As the capital of DRC, with a population of >17 million, identification in Kinshasa has further increased the complexity of the ongoing public health emergency and concerns regarding additional regional and/or international expansion of Clade I MPXV [11, 12]. The increasingly complex nature of MPXV transmission and association between sustained human transmission and dense populations, including sexual networks, has further broadened the definition of those populations considered at-risk for mpox. These at-risk populations include those who self-identify as men who have sex with men (MSM), sex workers, and people with frequent zoonotic contacts [13].

There is a critical need for rapid deployment of mpox diagnostics and medical countermeasures while also enabling healthcare access for at-risk populations in DRC [14]. Moreover, given the international expansion of Clade Ib to neighboring regions, including Burundi, Rwanda, Uganda, and Kenya, there is a critical need for community engagement and mobilization activities that are able to reach at-risk communities [3, 15–17]. This includes community-based research activities regarding vaccine uptake.

Here, we conducted a cross-sectional survey assessing mpox knowledge and sexual behaviours among MSM, sex workers and other at-risk populations in urban centers across three provinces in the DRC: Kinshasa, Kwango, and North Kivu. This investigation sought to assess mpox knowledge among populations at increased risk for mpox, including infection acquisition and transmission, while also evaluating high-risk sexual activities and health-seeking behaviors across these groups.

## METHODS

### Recruitment and Questionnaire Administration

The sampling methods have been described in detail previously [18]. In brief, between March 20, 2024, and August 25, 2024, we recruited 2,794 participants over 18 years old, self-identifying as either MSM and/or sex workers, and a general at-risk community group reporting neither sex work nor MSM activity. The recruitment was done in three urban study sites: Kinshasa in Kinshasa province, Kenge in Kwango province, and Goma in North Kivu province. Sex workers were identified among participants who indicated sex work as their primary or secondary occupation. Participants were categorized as MSM if they were classified as a man at birth and indicated ever having had sex with a male. MSM and sex worker networks were identified and accessed through facilities open to these communities or by peer educators who are connected to these communities. An additional at-risk population (ARP) group was formed of participants who self-defined as non-sex workers and non-MSM and were recruited by peer educators or at selected recruitment sites such as bars/clubs, major migration hubs and health centers. At-risk population participants included but were not limited to hunters, healthcare workers, retailers, butchers, or carpenters. Staff visited farms and abattoirs to enroll additional at-risk population members in close contact with animals–another route for possible mpox exposure.

At selected recruitment sites, participants were asked to complete in a tablet-assisted interview. The completed survey was stored on a secure server (SurveyCTO, Version 2.0, Dobility Inc.). Questions included demographic information, sexual activities, occupation, animal-related activities, and mpox knowledge. Key questions regarding Mpox knowledge and risk behaviors can be found in the attached supplemental materials (Supp. Table 1). Interviews were conducted by health-trained professionals in French or the local language, including Swahili, Kikongo, and Lingala.

**Table 1:**
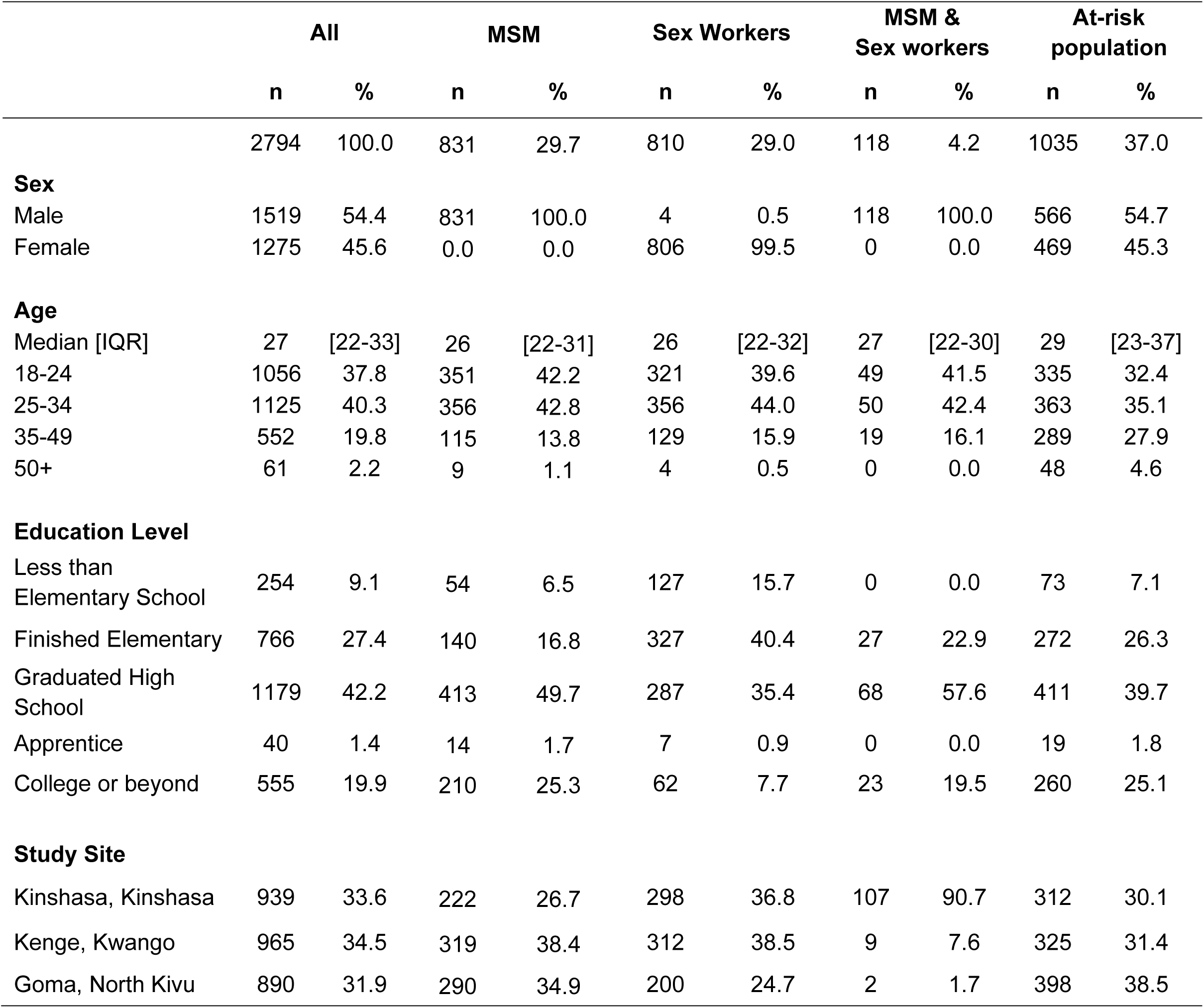
Demographic characteristics of participants.

### Ethics

Each participant was provided an informed consent prior to questionnaire completion. Approvals were provided by Institutional Review Boards of the University of California, Los Angeles (IRB#23-000676), University of Manitoba (HS25837) and the Kinshasa School of Public Health, University of Kinshasa (ESP/CE/161/2024).

### Data Analysis

Mpox knowledge was compared among MSM, sex workers, and the ARP, as well as across age groups and education levels. The three study sites were also compared against each other. Descriptive statistics were generated by calculating frequencies for demographic characteristics such as sex at birth, cohort type, age group, education level, and study sites. The chi-square or, where appropriate, Fisher’s exact test, was used to determine the association of categorical variables for bivariate analyses. Univariate analyses, specifically logistic regression models, were performed for variables with odds ratios (OR) and 95% confidence intervals (95% CI). Multivariate analyses included cohort type, age groups, education levels, and study sites, and results were considered significant at p-value < 0.05, with adjusted odds ratios (aOR) and 95% CI. All data analysis and figures were done using RStudio (R-4.2.2).

## RESULTS

### Demographic information

We recruited 2794 participants across three study sites in DRC, including 965 (34.5%) participants from Kenge, Kwango Province, 939 (33.6%) from Kinshasa, Kinshasa Province, and 890 (31.9%) from Goma, North Kivu Province. Among all participants, 1519 (54.4%) identified as male, and 1275 (45.6%) as female. The median age of participants was 27 years, with a total range between 18 and 86 years of age.

Among all participants, 810 (29.0%) self-identified as a sex worker as their primary or secondary occupation, with 806 (99.5%) of these respondents identified as female. Additionally, 831 (29.7%) of the participants self-identified as MSM. Of note, 118/2794 (4.2%) participants self-identified as both MSM and sex workers.

We also assessed education levels among study participants: 19.9% reported their terminal education level as college or beyond, 42.2% as completing high school, 27.4% as completing elementary school, and 9.1% as with less than an elementary school education (Table 1).

### Knowledge of mpox

Among all study participants, 6.1% (182/2794) had previously heard of mpox. This varied by population type: just 3.1% (25/810) of sex workers had heard of mpox the least of our population groups and significantly lower than both the MSM (7.7%, 64/831, p-value < 0.001) and ARP populations (8.8%, 91/1035, p-value < 0.001). Among participants that identified as both MSM and sex worker, 1.7% (2/118) had previously heard of mpox, which was significantly lower than ARP (p-value = 0.023). Mpox knowledge was significantly associated with age and education level (p-value < 0.001, respectively). Here, 10.7% (59/552) of participants aged 35-49 years had heard of mpox as compared to 6.7% (75/1125) of those aged 25-34 years (p-value = 0.03) and 4.0% (42/1056) of those 18-24 years (p-value < 0.001). The proportion of participants aged 50 years and older who had heard of mpox (9.8%, 6/61) was not significant compared to the other age groups (p-value > 0.05), Among those who reported less elementary school completion 0.8% (2/254) of participants reported having heard of mpox as compared to 16.0% (89/555) of participants with college education or beyond (p-value < 0.001). When considering geographic location, participants in Kinshasa reported less knowledge of mpox compared to those from Kenge (p-value < 0.001) or Goma (p-value < 0.001) (Figure 1). Logistic regression models for age groups and education levels indicated that those aged 35-49 years (aOR: 2.88, 95% CI: 1.83, 4.56) or 50 years and older (aOR: 2.89, 95% CI: 1.02, 7.05) were more likely to have heard of mpox compared to younger participants aged 18-24 years. Moreover, participants with a college degree or higher were more likely to have heard of mpox as compared to those who had not completed elementary school (aOR: 0.04, 95% CI: 0.007, 0.14), participants who finished elementary school (aOR: 0.13, 95% CI: 0.07, 0.22) and those who graduated from high school (aOR: 0.38, 95% CI: 0.27, 0.54) (Table 2).

**Figure 1:**
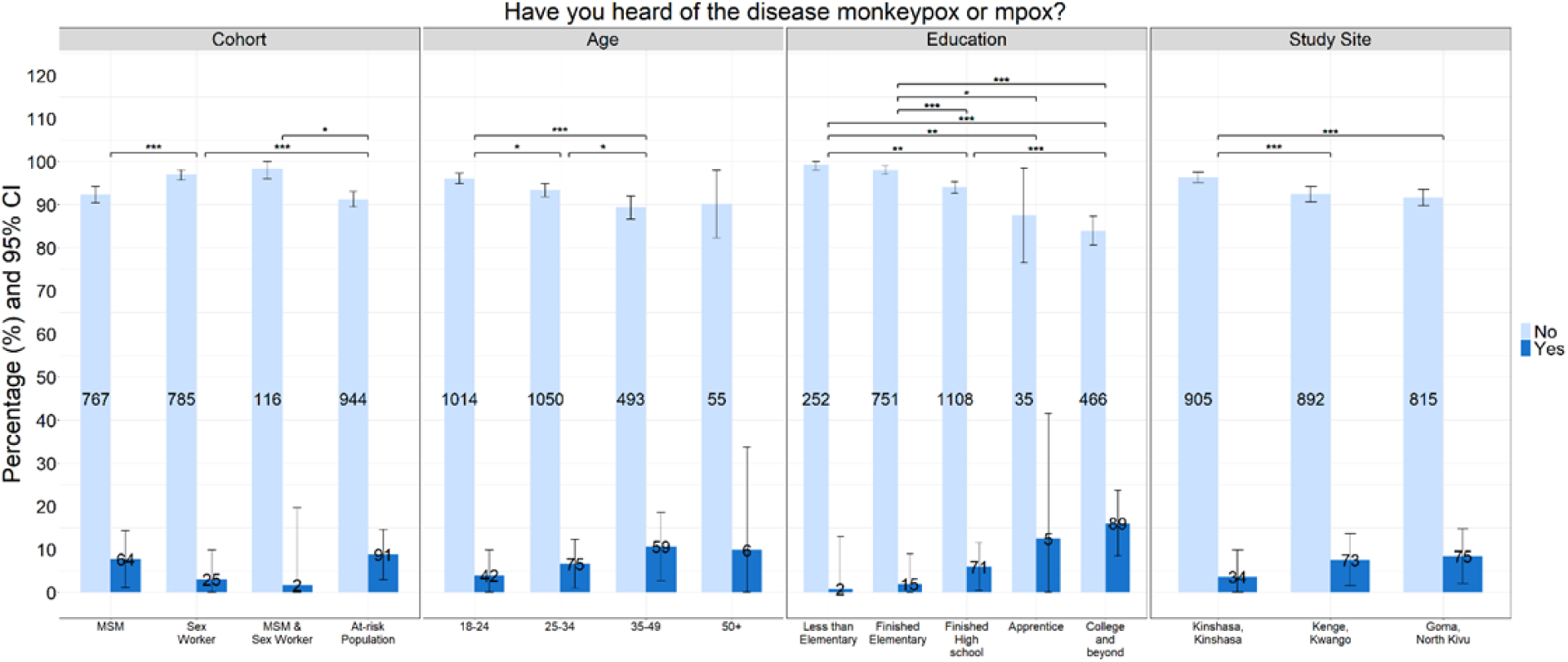
Knowledge of mpox. Questionnaire responses to “Have you heard of the disease monkeypox or mpox?” by different response groups. CI: confidence interval. *: p-value < 0.05, **: p-value < 0.01, ***: p-value < 0.001

**Table 2:**
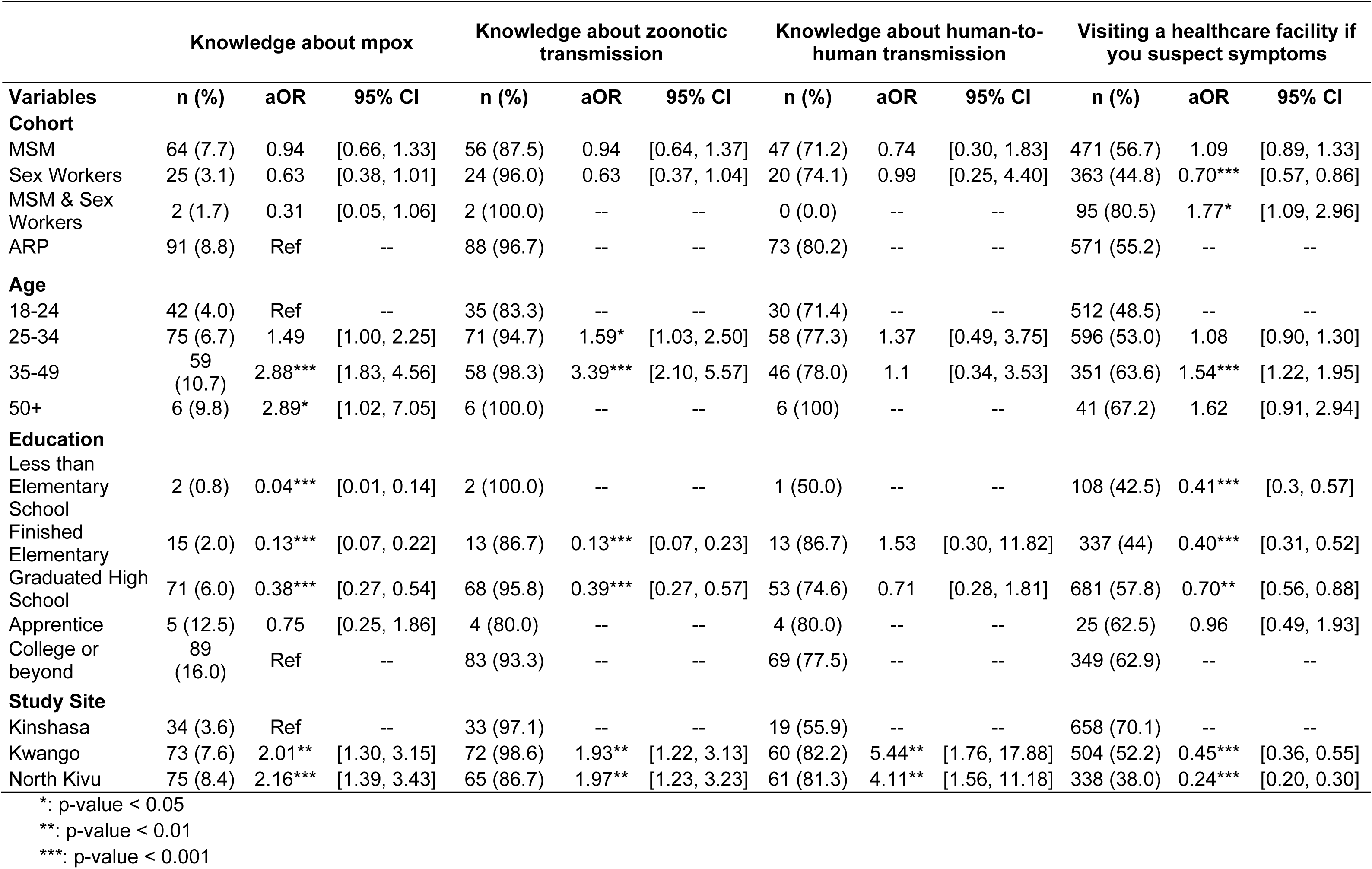
Odds of selecting an answer according to participants’ demographics.

### Monkeypox virus transmission knowledge

Among the 182 participants who had previously heard of mpox, 93.4% (170/182) of participants selected at least one zoonotic mode of transmission provided in the questionnaire, whereas 76.9% (140/182) of participants selected human-to-human transmission. Participants that identified as both MSM and a sex worker (n = 2), those aged 50 and older (n = 6) and participants with less than an elementary degree (n = 2) or apprentice (n = 5) were excluded from the regression analysis due to insufficient sample size. When compared to each other, we did not find any statistical significance between responses selected from MSM, sex workers, and ARP (p-value > 0.05) (Supp. Table 1).

We then assessed differences between mpox transmission knowledge and age, where the number of participants aged 18-24 years selecting a zoonotic transmission (35/42, 83.3%) was significantly lower than those ages 25-34 (p-value = 0.008) or 35-49 (p-value < 0.001). Our regression analysis demonstrated an association between knowledge of mpox transmission routes and age group—those aged 25-34 (aOR: 1.59, 95% CI: 1.03, 2.50) and 35-49 years (aOR: 3.39, 95% CI: 2.10, 5.57) were more likely to select a zoonotic route compared to those aged 18-24 (Table 2).

Geographically, 74.0% (54/73) of the participants from Kenge selected three MPXV transmission modes compared to 53.2% (41/77) of participants from Goma and 44.1% (15/34) of those from Kinshasa. Among the transmission mode options provided, participants from Kenge (aOR: 5.44, 95% CI: 1.76, 17.88) and Goma (aOR: 4.11, 95% CI: 1.56, 11.18) were more likely to select “human-to-human transmission” as compared to those from Kinshasa. Here, 55.9% (19/34) of participants from Kinshasa selected “human-to-human transmission” compared to 88.2% (30/34) of Kenge’s participants (p-value = 0.01) and 81.8% of participants from Goma (p-value = 0.02). Moreover, the number of participants selecting a zoonotic mode of transmission was significantly less from Kinshasa residents compared to Goma (86.7%, 65/75, p-value < 0.001) and those from Kenge (98.6%, 72/73, p-value < 0.001).

### Health-seeking behaviours among participants

Participants were also asked to select one action for the question: “Do you have any knowledge of how to recognize the signs of an STI and what action to take in the event of a suspicion?”. Of the total population, 53.7% (1500/2794) selected “I go to a healthcare facility,” 41.3% (1154/2794) selected “I treat myself with medication bought at the pharmacy,” and only 5.0% (139/2794) of participants selected “I do nothing”. More than half of the MSM (56.7%, 471/831) said they would go to a healthcare facility compared to 40.4% (336/831) of them saying they would treat themselves (p-value < 0.001) or do nothing (2.9%, 24/831; p-value < 0.001) (Figure 2). Among sex workers, we observed a similar trend, where 44.8% (363/810) would go to a healthcare facility, while 1.7% (14/810) would do nothing (p-value < 0.001) and 53.5% (433/810) self-treat with medication from a pharmacy (p-value < 0.001). As the age of the respondents increased, we observed an increase in reporting that they would seek care at a health facility (Table 2). Participants aged 35-49 reported being more likely to seek healthcare from a professional as compared to those aged 18-24 years (OR: 1.54, 95% CI: 1.22, 1.95). We observed similar trends by education level, where participants who completed high school (aOR: 0.70, 95% CI: 0.56, 0.88), had an elementary diploma (aOR: 0.40, 95% CI: 0.31, 0.52), and those with less than elementary school completion (aOR: 0.41, 95% CI: 0.3, 0.57), were less likely to go to a healthcare facility as compared to those with a college level or more.

**Figure 2:**
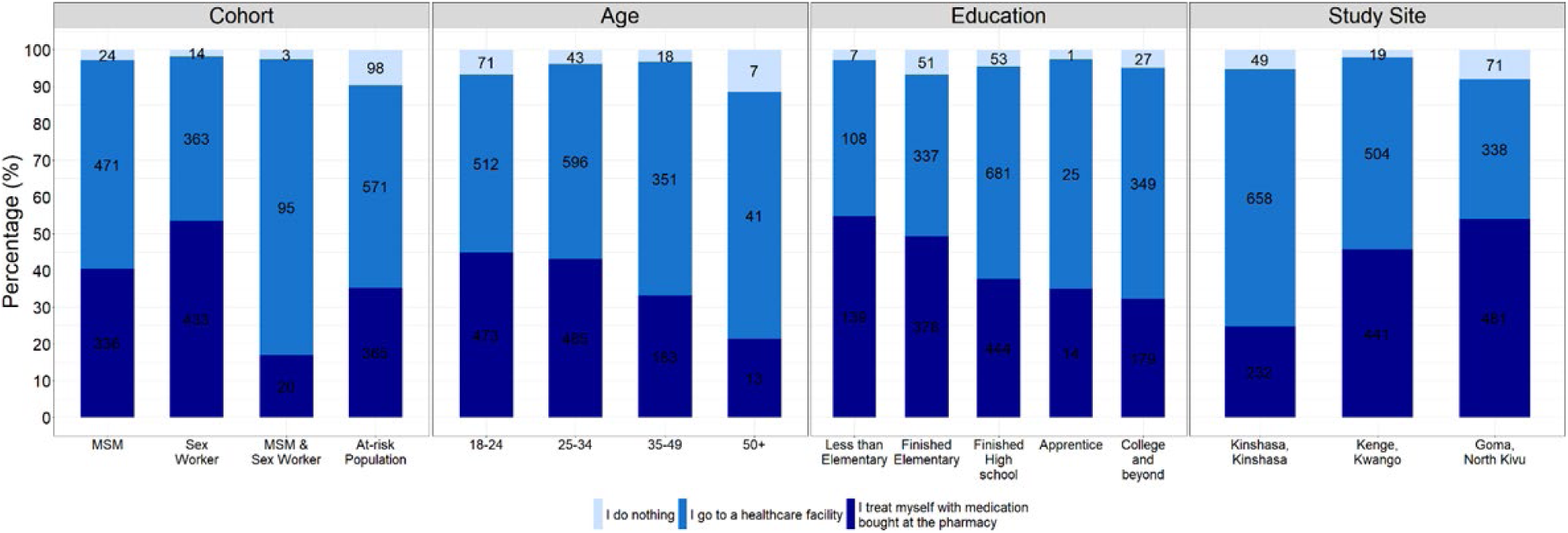
Health-seeking behaviours as reported by study participants. Percentage of participants for each course of action they would take in suspicion of an STI. CI: Confidence interval.

Furthermore, the results showed the opposite for the participants who would self-treat with medication bought at the pharmacy (Supp. Table 5). Indeed, participants with lower education, as those with less than elementary (aOR: 2.37, 95% CI: 1.70, 2.29) and those with elementary (aOR: 2.22, 95% CI: 1.72, 2.85) or high school completion (aOR: 1.37, 95% CI: 1.09, 1.73), were more likely to select that health-seeking behaviour as compared to participants with college education. When looking at geographic location, participants from Kenge (aOR: 0.45, 95% CI: 0.36, 0.55), and Goma (aOR: 0.24, 95% CI: 0.2, 0.3), were less likely to seek healthcare from professionals, but more prone to treat themselves (aOR: 2.64, 95% CI: 2.14, 3.27, and aOR: 4.06, 95% CI: 3.27, 5.07, respectively), as compared to participants from Kinshasa.

### Assessment of sexual behaviours and mpox risk

We assessed sexual behaviours of the participants considered to increase mpox acquisition risk by evaluating the self-reported frequency of five different types of sexual activities (Figure 3, Supp Tables 2 & 3). In the overall population, 59.4% (1661/2794) of all the participants said they had sexual relations or intimate contacts with more than one person in the prior 3 weeks. This was significantly higher among sex workers as compared to MSM participants, with 80.3% (745/810) of sex workers reporting multiple partners in the prior 3 weeks compared to 58.2% (484/831) of MSM participants (p-value < 0.001). Fewer respondents indicated having sex during travel, 33.7% (941/2794), as compared to other sexual activities provided in the questionnaire. Furthermore, 64.3% (1796/2794) of participants reported having transactional sex (including drugs, money, food or accommodations). When considering those that reported transactional sex participation among each group, a significantly higher percentage of those who identified as sex workers (84.7%, 786/810) answered yes as compared to those who identified as MSM (72.2%; 592/831, p-value < 0.001). Among all the population recruited, 57.8% (1616/2794) of participants reported having sex in clubs or bars. Stratified by population type, 79.9% (786/810) of sex workers and 59.1% (592/831) of MSM participants reported sex in clubs or bars (p-value < 0.001). Lastly, 1692/2794 (60.6%) participants said they ever had sex with anonymous partners, whereas most of the sex workers (80.8%, 750/810) reported this behaviour compared to MSM participants (59.6%, 495/831, p-value < 0.001). A significantly greater proportion of participants who self-identified as both MSM and sex workers reported having sex in exchange for goods (89.0%, 105/118) as compared to MSM (p-value < 0.001) or sex worker participants (p-value < 0.001) (Supp. Table 4). This was also similar when considering those participants who self-identified as both MSM and sex workers reporting having anonymous sex (84.7%, 100/118) compared to MSM (p-value < 0001) and sex workers (p-value = 0.002).

**Figure 3:**
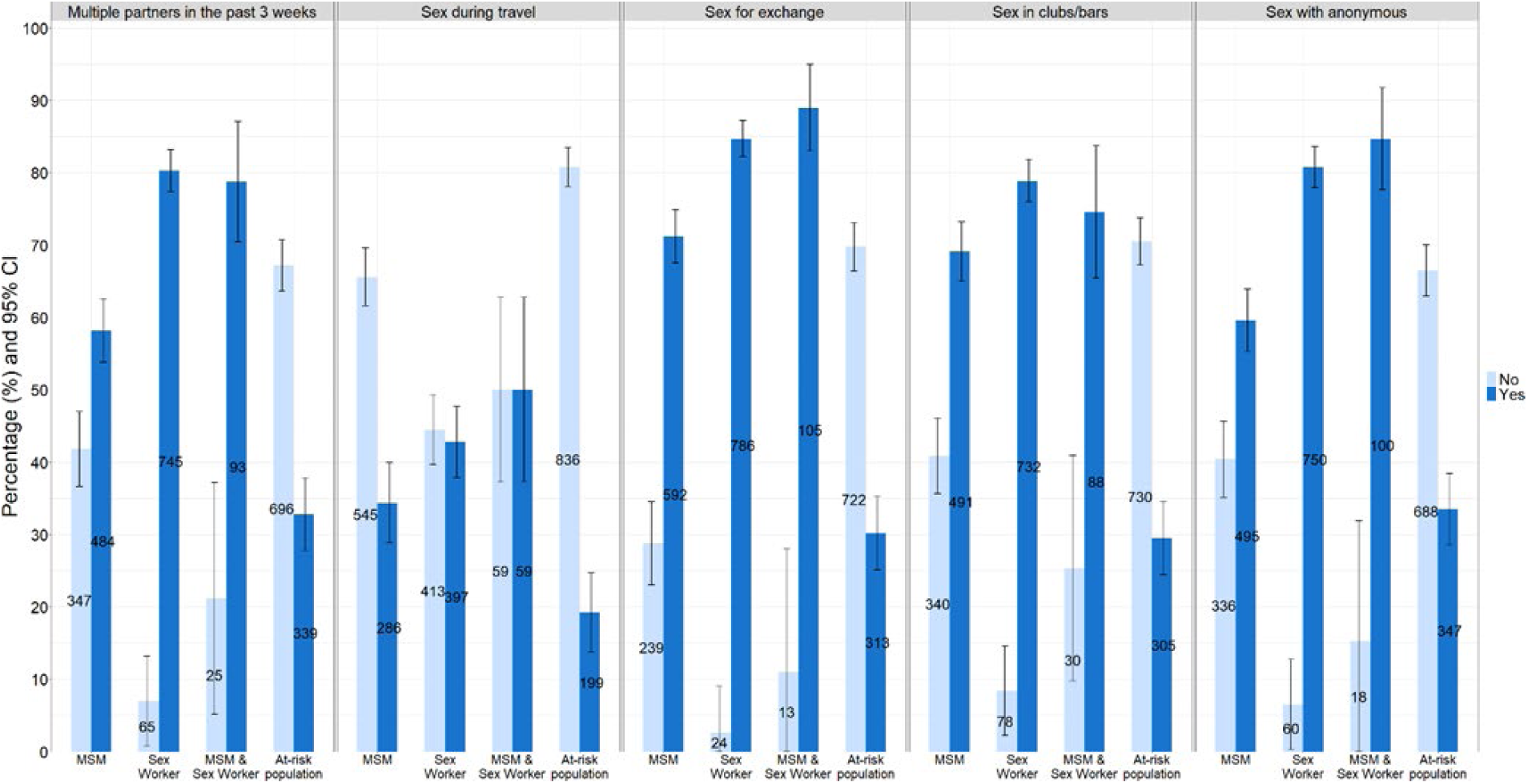
Assessment of sexual behaviours among study participants. Percentage of groups of participants depending on their sexual activities. CI: confidence.

## DISCUSSION

Our results show a suboptimal level of awareness of mpox among populations at increased risk for mpox in the DRC, even in the context of the current outbreak. Indeed, only 6.1% of participants said they had previously heard of mpox, this included 3.1% of sex workers and 7.7% of MSM. Because only two participants who identified as both MSM and sex workers reported knowing about mpox, we did not account for this in our analyses. These gaps in mpox knowledge among key populations further emphasize the need for improved mpox communication–with targeted public health outreach strategies specific to these groups. We report increased mpox knowledge among participants aged 24 years and older, as well as those with college or greater education. The role of participant age in mpox knowledge can be linked to the participant’s employment: older participants were often clinicians and health professionals, while most younger participants worked in an unrelated health field or were currently unemployed.

By study site, mpox knowledge was more common among respondents in Kenge and Goma than in Kinshasa. This could be explained by the higher education level of Goma participants, and the older population distribution in Kenge. It should also be noted that mpox has not been known to circulate historically in Kinshasa, with subclade Ia mpox first reported in the city in August 2023 and subclade Ib introduction in July 2024 [11, 12]. Given that the community engagement was made in a five-month timeframe, it should be appreciated that the variation in mpox knowledge found between participants in Kinshasa and other study locations may differ following the broader circulation of mpox within Kinshasa.

Among the participants who had reported hearing of mpox before, we further assessed their knowledge of the modes of transmission and populations at the highest risk for mpox; the majority of MSM and sex workers were able to identify modes of transmission. For all sub-populations, the least selected mode of transmission was “human-to-human”. This is problematic since human-to-human transmission has been documented among sex workers in South Kivu, DRC in 2024 [19], more globally among MSM during the clade IIb outbreak in 2022, and increasingly driving transmission in Kinshasa [13]. Moreover, the percentage of participants from Kinshasa selecting “human-to-human transmission” dropped to 55.9%, far less than the other study sites. This limited understanding of the transmission within the population needs to be addressed given the current co-circulation of subclade Ia and Ib in Kinshasa and reported sustained human-to-human transmission [13, 14]. When we looked at the answers to “Who is likely to contract mpox?”, the most selected response was “people living in high-risk areas”. This showed that people are aware of the risk in certain areas but do not necessarily realize that everyone can be at risk of contracting mpox. Our results in DRC contrast with mpox knowledge among Nigerian participants previously reported by Bakare *et al*. [20]. They reported a mpox knowledge among 38.3% of their participants; however, it should be appreciated that participants were recruited in healthcare facilities. Moreover, the Bakare *et al*. study demonstrated a good understanding of transmission routes and a high awareness about the perceived susceptibility of mpox among community members, with 58.8% of their participants aware that mpox can be transmitted during sexual intercourse. The contrasting results may demonstrate that considerable differences in mpox knowledge exist across geographic regions, even among endemic areas. It is also appreciated that our study focused recruitment on populations at increased risk for MPXV infection at convenient social locations instead of healthcare facilities.

As mpox transmission is increasingly linked to sexual contact, we wanted to determine the sexual risk behaviours of high-risk populations, including key population, in urban centers [21]. For each behaviour –sex in exchange for goods and services, multiple concurrent partners, etc.-we observed higher reported sexual activity among sex workers, followed by MSM, as compared to ARP respondents. However, while these participants showed to be at high risk for sexual transmissible diseases, the correlation between this and increased risk for mpox acquisition could not be determined. The lowest reported risk behaviour for each population was having sex during travel. However, the question was only focused on sexual activities during travel and no question was asked about general travel frequency. This could bias our analysis since participants could have answered “no” to this question only because they do not travel.

Finally, we sought to determine whether an individual would instead go to a healthcare facility, treat themselves or do nothing in the suspicion of an STI. The responses helped us understand if a participant was willing to seek help from a healthcare professional. This was related to understanding the behaviour of high-risk participants and whether they were open to receiving medication or vaccines as treatment or prevention for mpox. With the health belief model, we evaluated the perceived benefit of participants for seeking help in the suspicion of a sexual disease [22]. This model has been used for HIV (human immunodeficiency virus) prevention to understand factors associated with condom use and what recommendations could be made to prevent AIDS (acquired immunodeficiency syndrome) [23]. We showed that 59.8% of MSM and 49.2% of sex workers would consult a healthcare professional. Even with the stigma around their profession and/or their sexual activities, these groups are open to getting help from health professionals. This demonstrated that these individuals perceive the benefit of treating a sexual disease like STIs or mpox, by going to a healthcare facility, knowing they are at risk for stigma. The proportion of participants saying they would treat themselves decreased depending on their education level and age, where older people and those with higher education were more likely to go to a healthcare facility. This could be explained by the fact that older people had more knowledge and experience about STI and greater health-seeking knowledge. Similarly, people with higher education may be more informed about STIs, which correlates with the finding from Nigussie *et al*., who demonstrated that the knowledge of STIs increases for each year of study [24]. Furthermore, because 90% of healthcare is financed by private households in the DRC, older participants with higher education may have greater financial resources for treatment [25]. However, this might confound their answer, as they would be less likely to seek care for minor symptoms or illness if they must pay for it.

When considering the study location, participants from Kenge and Goma were more likely to treat themselves with medication from a pharmacist as compared to participants from Kinshasa. Oleffe *et al*. have shown that Congolese from Goma were more prone to purchase medicine at a pharmacy before seeking care from a healthcare professional due to better accessibility and lower costs [26]. Furthermore, a recent report from the DRC Ministry of Health showed that 90.0% of Kwango residents went to a public setting to seek healthcare, while 48.8% of Kinshasa residents went to a private healthcare facility [27]. While the perceived benefit of going to a healthcare facility was evaluated, questions about perceived barriers were not asked and would have been valuable to understand the different answers for the study cohort in our investigation.

Our study had limitations, which could affect the interpretation of our results. The sample size was focused on the different cohorts, i.e., MSM, sex workers and ARP, and the study sites. Further, age group and education level were not considered in the recruitment process, leading to skewed distributions of age and education level attainment groups. Moreover, for sexual activity-related questions, some questions had a time frame of three weeks or six months, followed by questions without a time frame which could have potentially impacted the interpretability by the participants.

In conclusion, this study highlights the important gaps in mpox knowledge among high-risk populations, high rates of high-risk sexual activities and limited understanding of mpox transmission routes. We demonstrated that the dearth of knowledge about mpox among key populations and frequent risk behaviours may contribute to the continued circulation of MPXV in urban centers, highlighting the need for targeted public health communication to at-risk cohorts. Further investigations are necessary to understand the accessibility of information and resources to protect them against diseases they can acquire in the course of their sexual activities. Moreover, paired with serological data, these data would help to elucidate the true burden of diseases among these key populations.

## CONTRIBUTORS

Conceptualization and study design: MH, NAH, AWR, PMK, JK. Field study coordination: MH, SM, LL, EHV, PKM, CK, MH, LL, RLS, NAH, AWR, PMK, JK. Data management: CL, MH, SM, YA, LL, PKM, EHV, TK, MK, CK, SL, NAH, AWR, PMK, JK. Writing - first draft: CL, MH, SM.

Writing - review and editing: CL, MH, SM, PKM, EHV, IIB, MC, GSG, MH, LL, SYS, LEH, NAH, AWR, PMK, JK. Supervision: MH, SM, NAH, AWR, PMK, JK. All authors contributed to the article and approved the final version of the manuscript.

## FUNDING

This work was funded by the International Mpox Research Consortium (IMReC) through funding from the Canadian Institutes of Health Research and International Development Research Centre (grant no. MRR-184813); Department of Defense, Defense Threat Reduction Agency, Monkeypox Threat Reduction Network (HDTRA1-21-1-0040); USDA Non-Assistance Cooperative Agreement #20230048; US NIAID/NIH grant number U01AI151799 through Center for Research in Emerging Infectious Disease-East and Central Africa (CREID-ECA); The content of the information does not necessarily reflect the position or the policy of the US federal government, and no official endorsement should be inferred.

## DECLARATIONS OF INTEREST

None declared.

## DATA SHARING

Due to the sensitive nature of our survey and the involvement of human participants we did not receive approval to collect raw participant identified data from research ethics boards/institutional review boards of the University of Manitoba, University of Kinshasa, and University of California, Los Angeles.

## Notes

### Competing Interest Statement

The authors have declared no competing interest.

### Summary of Updates

This revision is to add one additional author.

