## Supplemental tables for "Assessing mpox knowledge and sexual behaviours within high-risk populations in the Democratic Republic of the Congo"

**Supplemental Table 1**: Mpox knowledge questions and the possible responses

| Question | Responses |
|---|---|
| Have you heard of the disease monkeypox or mpox? | Yes<br>No |
| What are the modes of transmission of the mpox virus? | Direct contact with infected animals<br>Consumption of meat from infected animals<br>Human-to-human transmission |
| Do you have any knowledge of how to recognize the signs of an STI and what action to take in the event of suspicion? | I treat myself with medication bought at the pharmacy<br>I go to a health-care facility<br>I do nothing |

**Supplementary Table 2:** Comparison of mpox-specific answers of MSM and sex workers according to the at-risk population (ARP)

| Mpox-specific answers | Cohort | n | % | p-value |
|---|---|---|---|---|
| Direct contact with infected animals | MSM | 56 | 84.8 | 0.190 |
| | Sex Workers | 25 | 92.6 | 1.00 |
| | ARP | 84 | 92.3 | Ref |
| Consumption of meat from infected animals | MSM | 47 | 71.2 | 0.104 |
| | Sex Workers | 20 | 74.1 | 0.5540 |
| | ARP | 77 | 84.6 | Ref |
| Human-to-human transmission | MSM | 47 | 71.2 | 0.336 |
| | Sex Workers | 20 | 74.1 | 1.00 |
| | ARP | 73 | 80.2 | Ref |
| Anyone who has not been immunized | MSM | 46 | 69.7 | 0.069 |
| | Sex Workers | 19 | 70.4 | 0.371 |
| | ARP | 77 | 84.6 | Ref |
| People living in high-risk areas | MSM | 50 | 78.8 | 0.064 |
| | Sex Workers | 22 | 88.9 | 0.72 |
| | ARP | 82 | 90.1 | Ref |
| Healthcare workers in contact with confirmed cases | MSM | 40 | 60.6 | 0.001 |
| | Sex Workers | 17 | 63.0 | 0.074 |
| | ARP | 78 | 85.7 | Ref |

**Supplementary Table 3**: Comparison of sexual behavior of each group according to participant self-identified as MSM and Sex workers

| Sexual behaviour | Cohort | n | % | p-value |
|---|---|---|---|---|
| Multiple partners in the prior three weeks (n = 1661) | MSM | 484 | 58.2 | 0.0001 |
| | Sex Workers | 745 | 80.3 | 0.00004 |
| | MSM & Sex Worker | 93 | 78.8 | Ref |
| | ARP | 339 | 32.8 | 7.22e-22 |
| Sex during travel (n = 941) | MSM | 286 | 34.4 | 0.006 |
| | Sex Workers | 397 | 42.8 | 1.00 |
| | MSM & Sex Worker | 59 | 50.0 | Ref |
| | ARP | 199 | 19.2 | 1.79e-13 |
| Sex in exchange for goods (n = 1796) | MSM | 592 | 71.2 | 0.0003 |
| | Sex Workers | 786 | 84.7 | 0.0002 |
| | MSM & Sex Worker | 105 | 98.0 | Ref |
| | ARP | 313 | 29.5 | 1.72e-35 |
| Sex in clubs/bars (n = 1616) | MSM | 491 | 59.1 | 0.007 |
| | Sex Workers | 732 | 78.9 | 3.14e-06 |
| | MSM & Sex Worker | 88 | 74.6 | Ref |
| | ARP | 305 | 29.5 | 7.14e-22 |
| Sex with anonymous (n = 1692) | MSM | 495 | 59.6 | 7.26e-07 |
| | Sex Workers | 750 | 80.8 | 0.03 |
| | MSM & Sex Worker | 100 | 84.7 | Ref |
| | ARP | 347 | 33.5 | 1.67e-26 |

**Supplementary Table 4**: Comparison of sexual behaviour between MSM and Sex workers

| Sexual behaviour | MSM | | Sex Workers | | p-value |
|---|---|---|---|---|---|
| | n | % | n | % | |
| Multiple partners in the prior three weeks | 484 | 58.2 | 745 | 80.3 | 3.81e-55 |
| Sex during travel | 286 | 34.4 | 397 | 42.8 | 1.21e-08 |
| Sex in exchange for goods | 592 | 71.2 | 786 | 84.7 | 3.01e-45 |
| Sex in clubs/bars | 491 | 59.1 | 732 | 78.9 | 3.92e-47 |
| Sex with anonymous | 495 | 59.6 | 750 | 80.8 | 2.64e-54 |

**Supplementary 5**: Odds Ratios of selecting health-seeking behaviour according to the participants' demographics

| | | Visit to a healthcare facility | | | Self-treat with medication from the pharmacy | | | Do nothing | | |
|---|---|---|---|---|---|---|---|---|---|---|
| **Variables** | | **n (%)** | **aOR** | **95% CI** | **n (%)** | **aOR** | **95% CI** | **n (%)** | **aOR** | **95% CI** |
| **Cohort** | | | | | | | | | | |
| MSM | | 471 (56.7) | 1.09 | [0.89, 1.33] | 336 (40.4) | 1.24* | [1.02, 1.52] | 24 (2.9) | 0.28*** | [1.17, 0.44] |
| Sex Workers | | 363 (44.8) | 0.70*** | [0.57, 0.86] | 433 (53.5) | 2.06*** | [1.67, 2.54] | 14 (1.7) | 0.16*** | [0.08, 0.28] |
| MSM & Sex Workers | | 95 (80.5) | 1.77* | [1.09, 2.96] | 20 (16.9) | 0.78 | [0.45, 1.29] | 3 (2.5) | 0.22* | [0.05, 0.63] |
| ARP | Ref | 571 (55.2) | -- | -- | 365 (35.3) | -- | -- | 98 (9.5) | -- | -- |
| **Age** | | | | | | | | | | |
| 18-24 | Ref | 512 (48.5) | -- | -- | 473 (44.8) | -- | -- | 71 (6.7) | -- | -- |
| 25-34 | | 596 (53.0) | 1.08 | [0.90, 1.30] | 485 (43.1) | 1.00 | [0.83, 1.21] | 43 (3.8) | 0.69 | [0.46, 1.04] |
| 35-49 | | 351 (63.6) | 1.54*** | [1.22, 1.95] | 183 (33.2) | 0.76* | [0.60, 0.96] | 18 (3.3) | 0.46* | [0.25, 0.79] |
| 50+ | | 41 (67.2) | 1.62 | [0.91, 2.94] | 13 (21.3) | 0.50* | [0.25, 0.95] | 7 (11.5) | 1.08 | [0.42, 2.43] |
| **Education** | | | | | | | | | | |
| Less than Elementary School | | 108 (42.5) | 0.41*** | [0.3, 0.57] | 139 (54.7) | 2.37*** | [1.70, 2.29] | 7 (2.8) | 1.14 | [0.44, 2.64] |
| Finished Elementary | | 337 (44.0) | 0.40*** | [0.31, 0.52] | 378 (49.3) | 2.22*** | [1.72, 2.85] | 51 (6.7) | 1.67 | [1.01, 2.81] |

| | | | | | | | | | | |
|---|---|---|---|---|---|---|---|---|---|---|
| Graduated High School | | 681 (57.8) | 0.70** | [0.56, 0.88] | 444 (37.7) | 1.37** | [1.09, 1.73] | 53 (4.5) | 1.20 | [0.73, 1.99] |
| Apprentice | | 25 (62.5) | 0.96 | [0.49, 1.93] | 14 (35.0) | 1.11 | [0.54, 2.18] | 1 (2.5) | 0.70 | [0.04, 3.59] |
| College or beyond | Ref | 349 (62.9) | -- | -- | 179 (32.3) | -- | -- | 27 (4.9) | -- | -- |
| **Study Site** | | | | | | | | | | |
| Kinshasa | Ref | 658 (70.1) | -- | -- | 232 (24.7) | -- | -- | 49 (5.2) | -- | -- |
| Kwango | | 504 (52.2) | 0.45*** | [0.36, 0.55] | 441 (45.7) | 2.64*** | [2.14, 3.27] | 19 (2.0) | 0.44** | [0.25, 0.75] |
| North Kivu | | 338 (38.0) | 0.24*** | [0.20, 0.30] | 481 (54.0) | 4.06*** | [3.27, 5.07] | 71 (8.0) | 1.25 | [0.84, 1.88] |

*: p-value < 0.05
**: p-value < 0.01
***: p-value < 0.001
